# An exploratory application of failure mode and effects analysis to recruitment and retention challenges in a clinical trial

**DOI:** 10.64898/2025.12.06.25341655

**Authors:** Meichen Zhou, Fan Yang, Wanmei Wu, Xiaowen Zhou, Yan Li, Honglan Zhong

## Abstract

Clinical trials are a critical phase in the development of new drugs, and the recruitment of research participants is a pivotal prerequisite for the success of these trials. However, participants recruitment and retention often face numerous challenges, such as delayed recruitment progress, high rates of informed consent failure, and poor participants compliance, which significantly impact the timeline and quality of clinical trial projects. Through in-depth analysis of 4 core processes of participants recruitment and retention, this study explores the application of the Failure Mode and Effects Analysis (FMEA) method to identify four high-risk failure modes and three medium-high risk failure modes within the core processes of participants recruitment and retention. By implementing a multifaceted and systematically proactive approach, including leveraging established relationships with community providers, appointing dedicated full-time personnel, providing targeted incentives for both research staff and participants, and maintaining persistent communication, this exploratory study integrates preemptive risk identification and mitigation strategies aimed at participant recruitment and retention. This methodology seeks to establish a sustainable model for continuous improvement in clinical trial efficiency and participant engagement, focusing on the early detection and resolution of potential barriers.

## 1. Introduction

Clinical trials are essential for evaluating the efficacy and safety of new drugs. The design and implementation of clinical trials must ensure scientific rigor and data reliability, with the number and quality of participants being key to achieve these goals [1]. Participants recruitment and retention are cornerstones of successful clinical trials, as they directly influence statistical power, data quality, and the reliability of research conclusions. However, recruitment and retention remain notoriously challenging in clinical trials due to insufficient enrollment and high participant dropout [2]. Therefore, clinical investigators must dedicate significant energy throughout the trial, continuously maintaining their willingness and ability to remain engaged [3].

Failure mode and effects analysis(FMEA) is a systematic, proactive and teamwork-based tool for risks management, aimed to identify and assess the causes and effects of potential failure modes in a system, thereby preventing them from happening beforehand [4]. The possible failures were identified and analyzed by calculating the risk priority numbers (RPNs). An FMEA is performed by completing the following 5 steps: selection of processes to be assessed, construction of a multidisciplinary team, collection and classification of risk scores from each process, conduct of a risk analysis, and implementation of remedial actions for the failure modes and reanalysis to see if those actions are effective [5–7]. Although FEMA has been successfully used to inprove the quality of various clinical procedures in the hospital system, its application to the clinical trials system has been infrequent.

This manuscript integrates the FMEA into the management of participant recruitment and retention in a randomized clinical trial, to identify and evaluate potential failure modes in the recruitment and retention processes, including unexpected recruitment delays, unwillingness of engagement, non-compliance, and participant dropout. Potential consequences of poor participants recruitment or retention include delayed timelines and questionable results [8–10]. Corresponding improvement measures are proposed to mitigate those risks to optimize engagement and ensure implementation of the trial.

## 2. Material and Methods

### 2.1 Baseline FMEA

We conducted an FMEA into a phase Ⅲ, randomised controlled clinical trial conducted in a tuberculosis hospital, China, to evaluate the effects of strategies to improve recruitment of participants.To assess the baseline risk profile, an FMEA was performed from August 2023 to January 2024 by completing the following steps.

### 2.2 Formation of an FMEA focus team

An FMEA focus team was formulated, consisting of 8 multidisciplinary team members, collectively representing the quality improvement group, clinical research associates (CRAs), clinical research coordinators (CRCs), and clinical investigators. Additionally, a Project Leader from Parexel also joined the FMEA focus team to guide and assist the onsite FMEA processes. The team was responsible for the entire FMEA process, including the identification of failure modes, risk assessment, and the formulation and implementation of intervention measures. After assembling the team, educational and training lectures regarding FMEA were provided to increase the team’s knowledge about this project.

### 2.3 Identification of potential failure modes

The team developed a process map, which used flowcharts to illustrate the flow of a process(Fig 1). Through a multi-round, iterative evaluation process, the focus team selected 4 core processes that were deemed most prone to risk failure. Then, based on brainstorming and in-depth analysis, a total of 14 critical potential failure modes were systematically identified and prioritized for further analysis. The 4 selected core processes included: Identifying and reaching target audience, obtaining informed consent, screening and enrollment, Follow-up management.

**Fig 1.**
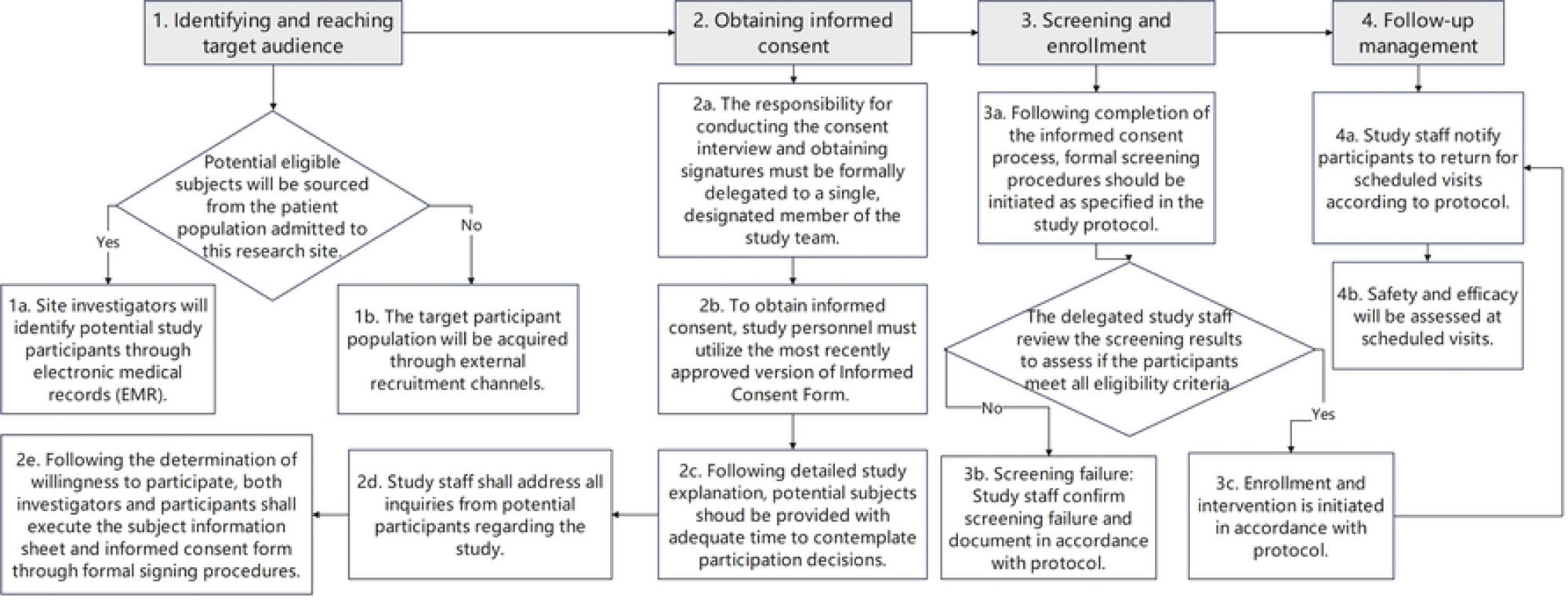
Core process map for FMEA in participants recruitment and retention.

### 2.4 Severity, Occurrence, and Detection Scoring

For each failure mode, its severity (S), occurrence (O), and detectability (D) were assessed, with a scoring range from 1 to 10. These scores were based on the degree of impact, frequency, and the probability of detection of each failure mode. Then, calculate the risk priority number (RPN = S × O × D) for each failure mode. Based on the percentile of RPN values (25%, 50%, 75%), failure modes were classified into four levels: “Low Risk, Medium-Low Risk, Medium-High Risk, and High Risk”, as shown in Table 1.

**Table 1.**
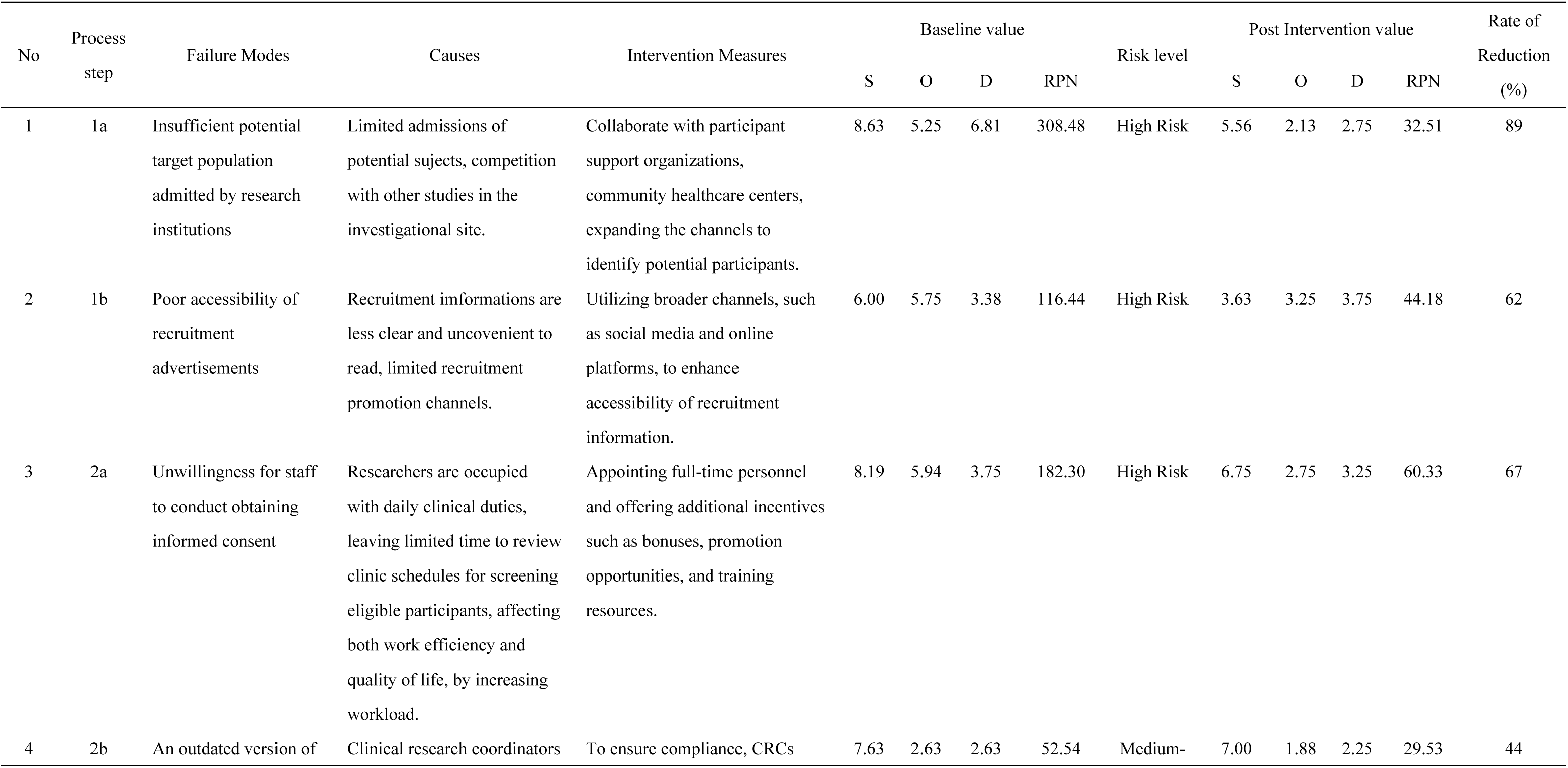
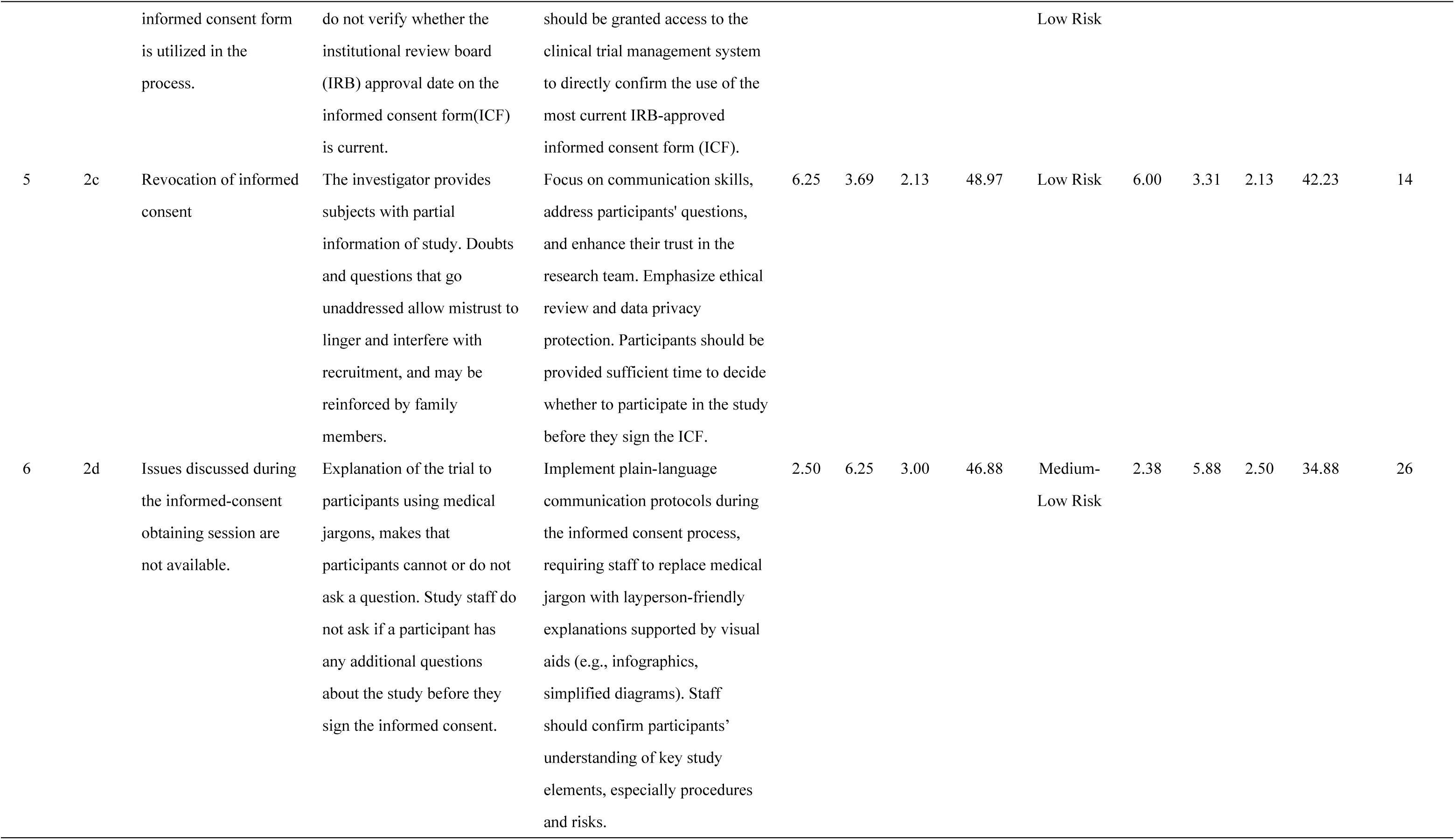
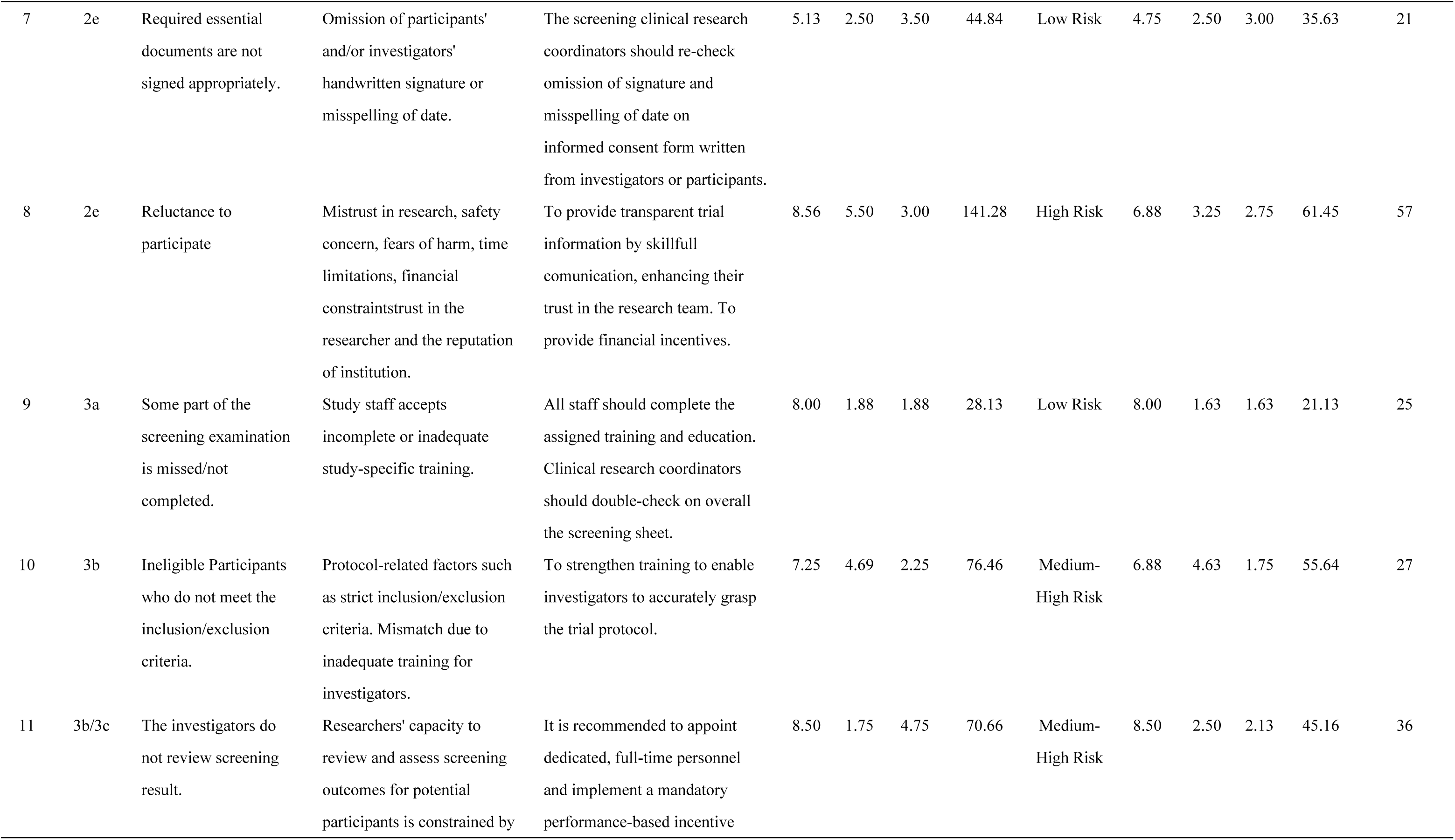
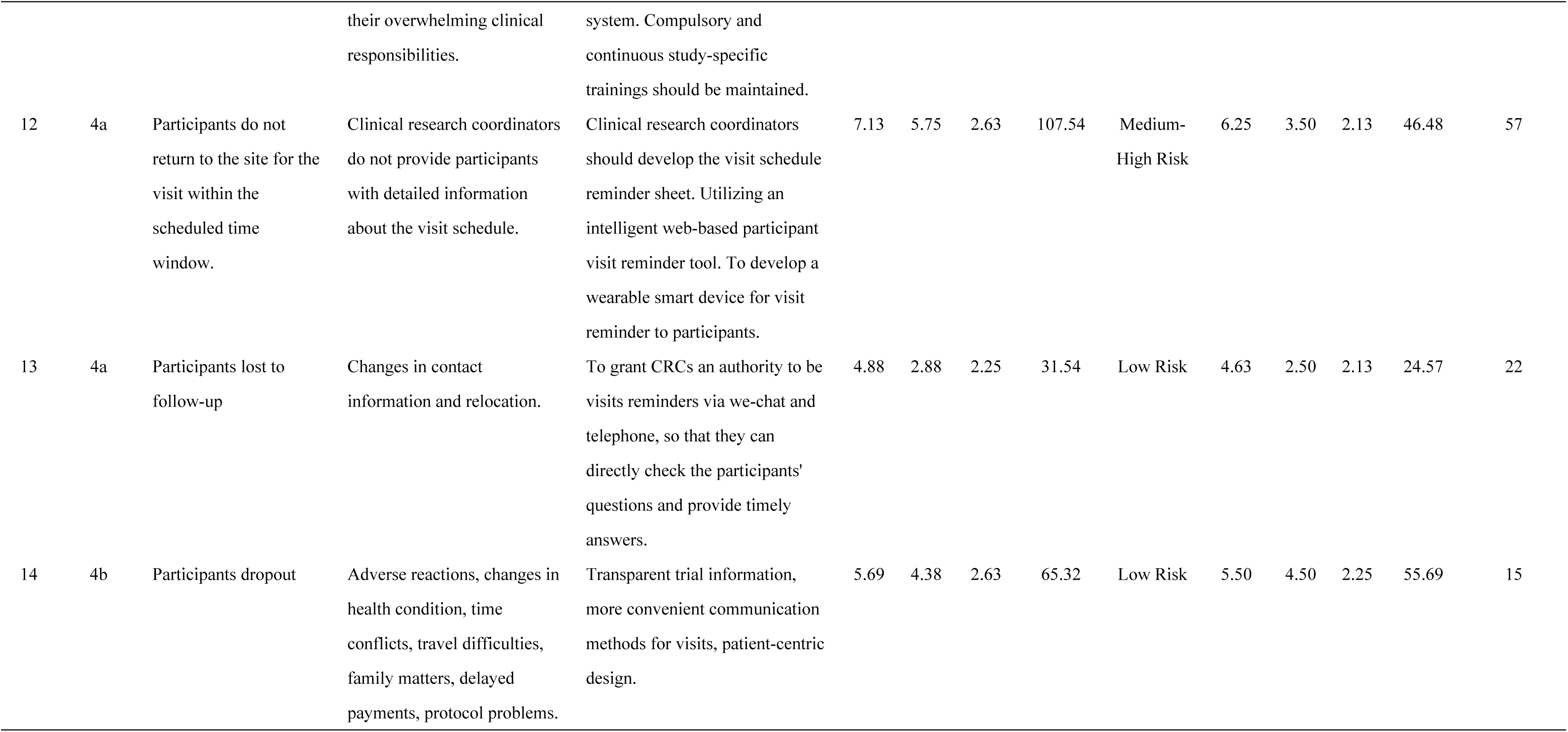
Assessment of RPNs for critical failure modes and effect analysis before and after intervention to improve recruitment and retention.

### 2.5 Intervention measures and effectiveness assessment

Based on the results of the baseline FMEA, targeted intervention measures were formulated and implemented for both “High Risk” (RPN>114) and “Medium-High Risk” (68<RPN<114) failure modes by focusing on reducing any of the S, O, and D scores. The effectiveness of the intervention was evaluated after three months in January 2024, by conducting a follow-up RPN assessment on the critical failure modes identified at baseline.

### 2.6 Ethics statement

This quality management study, which employed the Failure Mode and Effects Analysis (FMEA) methodology to optimize participants recruitment and retention processes, was conducted in accordance with the principles of the Declaration of Helsinki. This quality improvement project did not involve direct intervention with human subjects or the analysis of identifiable participant data. The study did not include minors.

## 3. Results

### 3.1 Initial RPNs

Based on 4 core processes, a total of 14 critical failure modes with many causes and effects were identified in all steps of the participants recruitment or retention (Table 1). As shown in Table 1, four main failure modes scored more than 114 in the initial RPN results and were identified as high risk and unacceptable failures, which was 28.6% of all failure modes. The four high-risk failure modes were associated with the first two core processes, namely identifying and reaching target audience, and obtaining informed consent. The three medium-high risk failure modes pertained to the latter two core processes: screening and enrollment, and follow-up management.

### 3.2 Effectiveness of targeted interventions on reducing risk priority numbers (RPNs)

For the 14 failure modes, this study formulated and implemented targeted intervention measures (Table 1). The re-evaluation results after three months showed that the risk priority numbers (RPNs) for these 14 failure modes all decreased, with reductions ranging from 14% to 89%, as detailed in Table 1. Following the intervention, the scoring factor for a specific failure mode was changed, primarily reflected by a slight increase in the occurrence score for “The investigators do not review screening result”, which rose from 1.75 to 2.50. A comparison of three-month averages before and after FMEA implementation showed an increase in monthly participants screening from 23 to 44, representing an 91% improvement. The overall screening failure rate was 24% (75/308). A total of 233 participants were randomised, of whom only 6 (3%) did not complete all scheduled visits.

## 4. Discussion

Recruitment and retention are challenges that involve much time and effort on the part of both clinicians and researchers. Identifying and reaching the target audience is the foremost critical element. Initially, researchers relied solely on electronic medical records (EMR) and limited recruitment advertisement channels to identify potential participants [11]. Although this method is straightforward, it has limited coverage, lower efficiency, and may fail to reach a diverse target population. Expanding the channels may be helpful to attract a broader audience and improve recruitment outcomes [12–13]. With the advancement of digital technologies, researchers can now utilize broader channels (such as social media and online registration platforms) for recruitment, thereby improving efficiency and sample diversity [14]. However, when adopting these new methods, it is also essential to pay attention to issues such as data privacy, ethical compliance, and technological accessibility to ensure the rigor and fairness of the research. Furthermore, The research staff can collaborate with participant support organizations, community healthcare centers, leveraging regular health education activities in the community to reach more potential research participants and enhance their trust in the research [15–16].

To invest additional time and cost is a critical barrier in research recruitment, especially for part-time investigators. Part-time investigators often need to balance their research with regular clinical practice or life responsibilities within limited time, and the time investment required for recruitment may further increase their burden [17–19]. Clinical Trial Site or teams can provide support, such as appointing full-time personnel, simplifying processes [20]. Offering additional incentives is an effective approach such as bonuses, promotion opportunities, and training resources for those dedicated investigators, based on their contribution to clinical research. The observed increase in the occurrence score for “The investigators do not review screening result” (from 1.75 to 2.50) may be attributed to inclusion of rotating dedicated investigators who received incomplete or inconsistent training, leading to perceived elevations in the risk of this particular failure mode among some team members. This can be intervented and improved by continuous reinforcement training.

Participants have concerns about engaging to the trial, including mistrust in the research, safety concerns, fears of harm, time constraints, inconvenience caused by frequent visits, financial limitations, and trust in the investigator and the reputation of the institution [21–23]. These factors lead to their reluctance to participate. Addressing this issue requires multifaceted measures. Investigators should provide transparent trial information, emphasize ethical review and data privacy protection, and demonstrate the professionalism of the research team and the site reputation. Investigators need to focus on communication skills, address participants’ questions, and enhance their trust in the research team. Providing financial incentives or medical resource support can strengthen willingness to engage. Utilizing digital visit communication platforms can reduce the time and cost of face-to-face visits while offering participants more convenient communication methods, which is beneficial for participant retention.

Participants may fail to return to the site within the scheduled visit window, which could be attributable to insufficient communication from clinical research coordinators regarding visit details. To mitigate this issue, it is recommended that coordinators develop a standardized visit schedule reminder sheet [24]. Furthermore, the implementation of an intelligent web-based reminder system could enhance adherence [25]. Additionally, the development of wearable smart devices designed to deliver visit reminders may offer a promising approach to further support participant compliance.

## 5. Conclusion

In recent years, FMEA has been increasingly applicated in medical field, demonstrating broad prospects. FMEA has not only played an important role in enhancing medical safety but has also contributed to improve risk management in medical processes. Furthermore, FMEA has shown significant effectiveness in risks management for human subjects in clinical research.

Recruitment and retention are vital challenges which can make or break clinical trials. Effective strategies include using multi-pronged recruitment avenues and strategy, cooperating with community healthcare centers that have been established trusted relationships with potential participants, employing full-time study staff, helping participants process and overcome ambivalence by skillful communication, increasing incentives for study staff and community providers contributed to recruitment, and optimizing the compensation disbursement process. Those strategies can provide an overall positive experience for both participants and study staff. FMEA offers a systematic framework for identifying and mitigating potential risks in clinical trial processes. Our FMEA focus team are extending the application of FMEA to other core trial operations, demonstrating its utility as a scalable and adaptable tool for quality improvement in randomized controlled trials. This approach represents a proactive methodology for risk management and process optimization aimed at reinforcing trial robustness and reliability.

## Data Availability

All relevant data are within the manuscript and its Supporting Information files.

## Author Contributions

**Conceptualization:** Meichen Zhou, Fan Yang, Wanmei Wu.

**Formal analysis:** Meichen Zhou, Fan Yang.

**Supervision:** Meichen Zhou, Wanmei Wu, Xiaowen Zhou, Yan Li, Honglan Zhong.

**Writing – original draft:** Meichen Zhou, Fan Yang, Wanmei Wu.

**Writing – review & editing:** Meichen Zhou, Fan Yang, Wanmei Wu, Xiaowen Zhou, Yan Li, Honglan Zhong.

## Acknowledgments

We are sincerely thankful for the contributors’ invaluable insights and steadfast support to the accomplishment of this research endeavor.

